# Fair Reinforcement Learning for Maternal Sepsis Treatment

**DOI:** 10.1101/2022.08.09.22278582

**Authors:** Siân Carey, Ciarán McInerney, Tom Lawton, Ibrahim Habli, Owen Johnson, Leila Fahel, Alwyn Kotzé, Marc de Kamps

## Abstract

**Objectives:** Reinforcement Learning is a branch of artificial intelligence (AI) which has the potential to support significant improvement in patient care. There is concern that such approaches may reinforce existing biases within patient groups. Understanding discrimination in AI models is important for building trust and ensuring fair and safe use. We explore the fairness of a published reinforcement learning model, used to suggest drug dosages for sepsis treatment of patients in critical care, on whether it safe to use with maternal sepsis patients.

**Methods:** We evaluate the current model using by a) comparing the results for a group of patients with maternal sepsis against a matched control group and b) using random forests to explore feature importance in the model.

**Results:** Our results show that the original clinicians’ decisions and model suggestions were similar across cohorts. Our feature importance ranking shows high variance for many of the features.

**Discussion:** In medical settings, different subgroups may have specific clinical needs and require different treatment however, in the absence of a clinical consensus on the most appropriate treatment, AI algorithms that give consistent treatment to patients regardless of subgroup could be judged as the safest and fairest option.

**Conclusion:** Our experiments showed that the evaluated model gave the same treatment to maternal and non-maternal sepsis patients. The methods developed for evaluating fair reinforcement learning may be more generally applicable to understanding how clinical AI tools can be used for safely and fairly.

**What is already known on this topic:** The use of reinforcement learning to suggest drug dosages for sepsis patients in critical care is a well-researched area, with multiple open-source models available. It has not previously been considered whether these models can be used on maternal sepsis patients.

**What this study adds:** The model studied behaves consistently on maternal and non-maternal sepsis patients, and appears to suggest an increased use of vasopressors compared with historical actions.

**How this study might affect research, practice or policy:** This study shows that it is possible to design models which are consistent across maternal and non-maternal sepsis patients, suggesting that a single model may be appropriate across a variety of patients with sepsis.

## INTRODUCTION

Sepsis is a life-threatening organ dysfunction caused by a patient’s response to an infection [1]. Sepsis is a leading cause of mortality across the world with an estimated 5.3 million deaths annually [2] and was recognised in 2017 as a global health priority by the World Health Organisation [3]. Sepsis patients are commonly treated using vasopressors and intravenous (IV) fluids, and despite the high mortality rate, there is currently no optimal plan for the administration these in the treatment of sepsis patients [4].

When sepsis develops during pregnancy, during birth, after birth, or following an abortion, it is categorised as maternal sepsis [5]. In 2019, sepsis accounted for a quarter of maternal deaths in the United Kingdom [6] and was the second highest cause of pregnancy related death in the US between 2014 and 2017 [7]. In low- and middle-income countries, deaths from maternal sepsis are reported at an even higher rate [8]. There are no internationally recognised diagnostic criteria specific to sepsis in pregnant patients [9] and many early signs of sepsis can be dismissed as common pregnancy symptoms. Due to this, clinicians are frequently more careful with pregnant patients and suspected sepsis, and the UK national criteria for sepsis are more stringent for pregnant patients than for non-pregnant patients [10].

Sepsis is currently treated in a critical care setting with the use of both IV fluids and vasopressors. Multiple vasopressor and IV fluid treatment plans have been tested with a range of results in terms of patient mortality [11]. In our evaluation, we focus on the safety concerns around the rate of change of vasopressor dosages [4]. There is disagreement over whether maternal sepsis should be treated in the same way as it is for the rest of the population [6]. Anecdotal evidence for clinicians in the UK NHS suggest that although vasopressors would be expected to have similar effects in pregnant and non-pregnant patients, they are generally used less in pregnancy likely due to fear of effect on the baby or due to capabilities of the ward treating the patient.

The optimal clinical pathway for sepsis treatment has not yet been found [4]. With the increasing use of electronic health records, research has been conducted using historic data into finding a gold standard treatment plan. In previous work [12–14] reinforcement learning, a branch of artificial intelligence (AI), has been used with electronic healthcare records to find an optimal dosage for IV and vasopressor treatment in sepsis. Further work has also been done on the safety of this work. Notably, Jia et al. [15], modified Raghu et al.’s [13] work for safety concerns by reducing the number of potentially dangerous large jumps in vasopressor dosage.

One of the aspects of ensuring patient safety is to confirm that protected characteristic groups receive outcomes that are as safe as everyone else and therefore can be considered “fair”. One such protected characteristic is pregnancy and maternity, which has historically seen a lack of information and research [16,17]. Therefore, we evaluate the recommended dosage policy with respect to maternal sepsis patients.

Most literature at the intersection of medical AI and identifying fairness concerns involves testing a model at the end of its creation [18,19]. Often this is done using what are known as fairness notions, such as statistical parity or equalised odds [20], which are statistical tests that show if particular standards are met in the results. Each fairness notion has particular requirements of the model or data, such as the availability of a ground truth or the cost of miss-classification [21]. However, due to the nature of reinforcement learning models there are not currently any notions suitable for use [20]. Alternative methods for testing discrimination include visualisation and interpretation of results and investigation of the original data.

## METHODS

### Dataset Description and Pre-processing

We used the MIMIC-III open access critical care database consisting of 53,423 distinct hospital admissions for patients admitted to a tertiary hospital in Boston, USA [22,23]. As previously done by Jia et al. [15] and Raghu et al. [13], for training of the model all participants met the sepsis-3 criteria [1] at some point during their time in critical care. Participants were excluded if: they were under the age of 18; their IV fluids or vasopressors were not recorded; treatment was withdrawn; or if they had extreme fluid intake. The latter reasoning was due to the likelihood of extreme fluid intake representing an incorrect chart rather than real events. There were 47 features recorded for each participant, for every 4 hours they were in the intensive care unit (ICU), as in the previously published work [13,15]. More information can be found in the addendum to Raghu et al. [13], whose previously published criteria we are reusing.

As we were evaluating for maternal sepsis, the test group included all participants that were recorded as having one of the pregnancy related ICD-9 codes (630-679 and 760-779) [24]. The test group consisted of all relevant individuals found in the data, which gave a cohort of 74 patients. The control group, matched within 5 years of age and sex and randomly selected from the previous participants, was created consisting of 370 individuals.

A descriptive analysis of the cohorts is in Table 1, including Sequential Organ Failure Assessment (SOFA) scores and 90-day mortality. The SOFA score is a measurement of organ failure, where higher numbers indicate poorer outcomes. The 90-day mortality is a binary result for each patient, recorded as a 0 if the patient survives for 90 days post hospital and 1 otherwise.

**Table 1:** Details about the cohort used in this report.

|  | Pregnancy Cohort | Control Cohort |
| --- | --- | --- |
| Unique ICU admissions | 74 | 370 |
| Mean age | 32.4 | 32.4 |
| Mean initial SOFA score | 6.7 | 6.9 |
| Mean 90 day mortality | 0.3 | 0.3 |

### Reinforcement Learning

We recreated the double deep duelling Q-network model presented in Jia et al. [15] to learn the optimal dosages for vasopressor and IV fluid given to sepsis patients in critical care.

Reinforcement Learning involves an agent learning the best approach to deal with a situation by receiving feedback (in the form of rewards) from how a model environment (state) responds to actions the agent takes. A Q-network model takes feedback from multiple future possibilities predicted by the model, rather than just considering the rewards at the immediate next stage [25]. A deep Q-network model uses a neural network to find the best action to take, that is the action from which it receives the highest reward. A double deep Q-network model adds a second neural network to act as the target, so that the action to be tested is chosen by the target neural network instead of a table. This improves the stability of the learning. A double deep duelling Q-network allows for separation to consider whether the resulting state is a positive outcome because the action was well chosen, or because the initial state was already positive.

This model does not learn from the clinicians actions directly, as the vasopressor and IV fluid dosages given by the clinicians are not features in the model, but from observations taken from the patient. However, there will be some indirect learning as it can only see the consequences of what did happen rather than what might have happened.

This model was created on the full sepsis cohort with a split of 85:5:15 for the respective training, validation and test sets. Each patients’ ICU records were broken into four-hour blocks, with one block representing one state for the model. The intermediate rewards use 1) SOFA score, and 2) arterial lactate, which is the level of lactate in the arterial blood. Combined these rewards range between +10 and −10. The final rewards are based on the 90-day mortality for each patient, with +15 for survival and −15 otherwise. At each point these rewards are combined, considering how soon each reward would happen, to decide the overall reward. The model recreated here is the same as that presented in Jia et al. [15]. Code was written in Python and can be found at https://github.com/SianC/Maternal_Sepsis.

### Visual Interpretations

The first of the two ways we investigated the model is by comparing the predictions made with the original decisions recorded in the data set, for each of the cohorts. There are two cohorts that we use to test the model: pregnant sepsis patients, and a matched control group. We focus on the suggested vasopressor dosages, and present both heat maps and bar graphs that visualise the differences between the model suggestions and original decisions, as well as between the different cohort suggestions and original decisions.

Anecdotally, it is widely debated between clinicians as to whether pregnant patients with sepsis should be treated differently to non-pregnant patients with sepsis, however the presumption is that they should be treated similarly unless evidence suggests otherwise. Therefore, we judged fairness partially on how similar the suggestions are for the different subgroups.

### Feature Importance

Some features have different baselines in pregnant and non-pregnant patients and, therefore, if these features are deemed to have a higher importance by the model diagnostics, then it is more likely that the model will not work as well on maternal sepsis. We used a random forest classifier to order the features by their importance to the model, that is their effect on the output of the model. These importance levels will then be evaluated for their relation to pregnancy and the likelihood of different baseline results being normal. For example, it is well known, that white blood cell count is higher in pregnant patients than non-pregnant patients [26].

We created ten models using the same framework and then found the feature importance for each model. On this basis we found an average importance score, and range, for each feature.

## RESULTS

### Data Description

Table 1 shows a breakdown of the age, sex, 90-day mortality and SOFA score for each cohort.

### Visual Interpretations

Figure 1 shows heat maps that describe the results from testing the model on the pregnant and control groups. Each vertical set of two heat maps represents a different cohort, and each horizontal set of heat maps represents a different policy: the top row represents the original clinician’s policy; the bottom row represents the Reinforcement Learning model’s policy. The numbers across the bottom of the heat map represent categories of vasopressor dose given, where 0 is no vasopressor and 4 is greater than 0.45 mcg/kg/min of vasopressor. Similarly, the column represents categories of IV fluid dose, with the same ranking. The darker a square is on the heat map, the more often that vasopressor-IV fluid dose pairing was given (in the case of clinicians) or suggested (in the case of the model) to a patient.

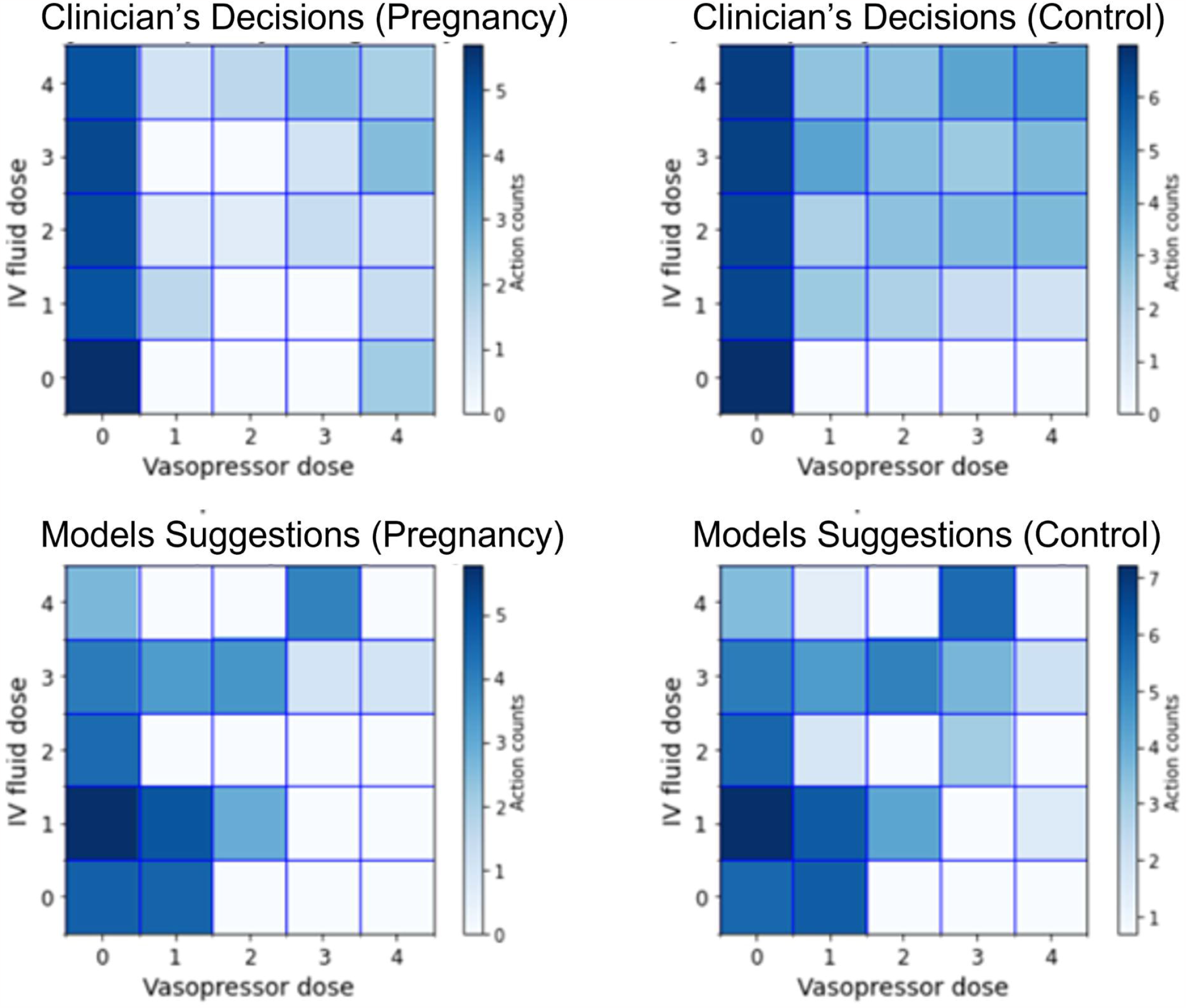

Figure 2 shows the same results with a focus on vasopressor dose. Each bar shows the percentage of patients from the relevant cohort and relevant decision maker who had that range of vasopressor dose. For example, the model gave 20% of the control cohort a dose of 0.

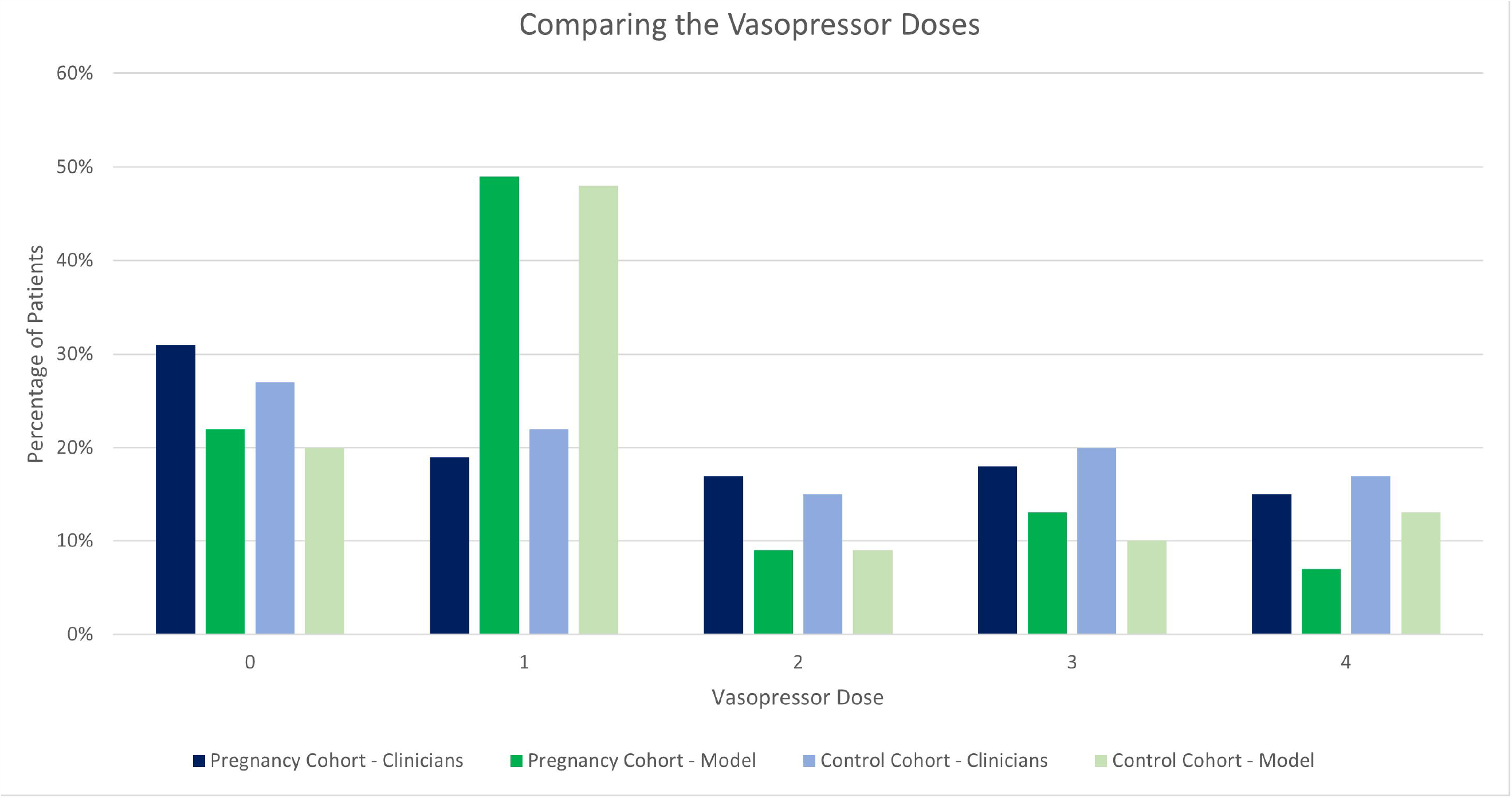

### Feature Importance

The left-hand plot in Figure 3 shows the feature importance for the model created to produce the same results as the original clinicians and is shown as a benchmark. The right-hand plot in Figure 3 shows the mean average feature importance for the deep Reinforcement Learning model presented in this paper. The error bars show the minimum and maximum importance score each feature received during the ten model runs. A high score indicates a higher level of importance.

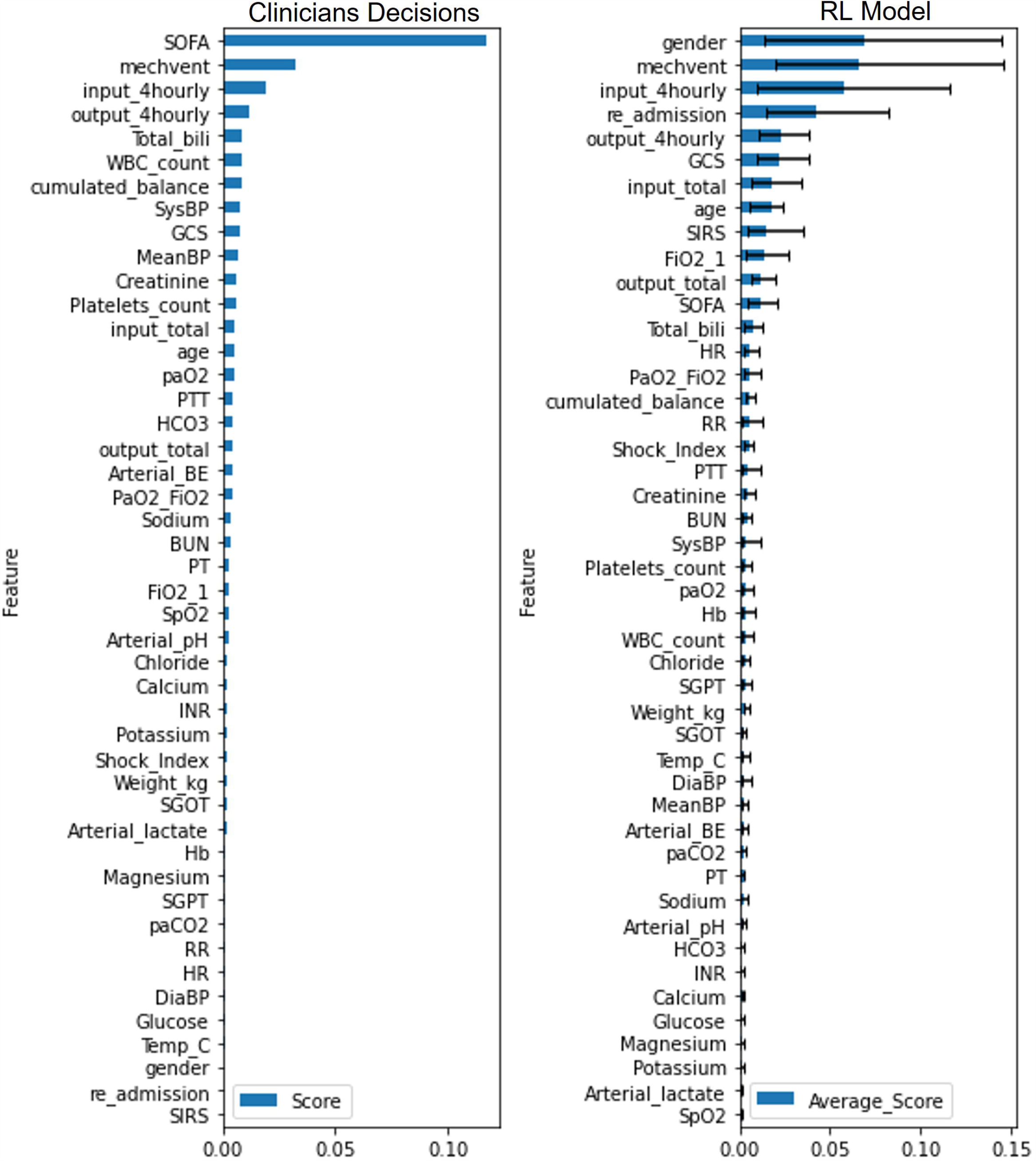

## DISCUSSION

The main novel contribution of this report is its study of maternal sepsis in relation to wider sepsis drug prediction models, as this is an area lacking from current literature. We explain the results and set them in context below.

### Visual Interpretations

These results show that there is less difference in model-suggested treatment than in the clinician’s decisions. The top row of Figure 1 shows the clinician’s decisions for the two cohorts, for both IV fluid and vasopressor. The main difference between the two cohorts is a higher amount of vasopressor being given to the control cohort, although this difference is slight. In comparison, the bottom row showing the models suggestions have less difference between the two graphs. The debate of how to treat maternal sepsis is still active within the medical field and some believe, as previously mentioned, that the safe and fair choice is to treat pregnant and non-pregnant patients the same until further research has been done. This model meets these expectations.

The difference between the clinician’s decisions and model suggestions are visibly similar. In Figure 2 the percentage of cohorts getting a certain vasopressor dosage can be directly compared. This also shows the similarities between the suggestions for the two cohorts seen above. The largest difference between the model and clinician is the number of patients suggested a vasopressor dose of “1” by the model. This is likely due to the limited cohort sizes, leading to a small lead appearing much larger when percentages are considered.

MIMIC-III was collected between 2001 and 2012. In the decade since then normal treatment has already moved in the direction suggested by the model, with higher amounts of vasopressor being used. However, as vasopressors can only be given on critical care wards within the UK this increase may not be as much as expected.

### Feature Importance

The size and similarity of the error bars on the second graph of Figure 3 indicate how unstable this method is for evaluating a model. Although not all of the error bars are as wide as those featured at the top of the graph, the similarity in width among features lower on the graph suggests that any feature between Sp02 (oxygen level) and HR (heart rate) could swap with any other. Furthermore, those at the top between gender and SOFA score also have overlapping bars.

It is notable that those features with the largest mean average also have the greatest variance between model runs. This is likely due to the variability within the model. Within a single model there is the possibility that one feature will be used more than others. As the top range of features all show similar mean average scores, it is understandable that they all experience being the primary feature when enough models are run. It would not be possible to make any reasonable conclusion using this method and expect it to be reproducible, other than this not being a useful evaluation tool in this situation.

## CONCLUSION

Maternal sepsis is a life-threatening condition that can occur during pregnancy, birthing or abortion and is among the leading causes of maternal mortality across the world. There are multiple published models that look at the prediction of sepsis drug dosages, however none of them focus on maternal sepsis. The model recreated here, originally presented by Jia et al. [15], used Reinforcement Learning to recommend the amount of IV fluid and vasopressor that sepsis patients should receive in critical care. We have shown that this model suggests similar amounts of vasopressor to maternal and non-maternal patients, however due to the size of the cohorts involved, it is not possible to take this as a strong conclusion. This implies that the evaluated model is safe and fair to use on maternal sepsis patients, however further data would be required to strengthen this conclusion.

### Limitations

The most significant limitations of this work relate to the size and provenance of this data. The primary cohort of pregnant patients contains only 74 records, which is too small a number to draw strong conclusions from. Additionally, the data is drawn from historic data from a single US hospital, and as such is unlikely to be ethnically (due to location) or class (due to the US healthcare system) diverse. This means that intersectional research has not been done. Finally, this data only consists of patients who spent time in intensive care, and does not take into account patients that did not enter intensive care.

### Future Work

Continuing this work with larger and more representative data sets would allow for more trustworthy predictions and would certainly need to occur before results could be regarded as clinically significant. Additionally, the data sets must be representative specifically for the population where the model is to be put into use.

We have deemed that the model studied here behaves consistently on pregnant and non-pregnant patients, as there are no significant differences in results between the cohorts in the study and no risky features ranking highly in the feature importance list. However, this does not mean that it is not important to continue this area of study, as any models that are considered for use in clinical practice should be tested like this to ensure that there are no inherent biases in the system. The methods developed for evaluating fair reinforcement learning may be more generally applicable to understanding how AI tools can be used for safe and fair clinical use.

## Data Availability

All data produced in the present work are contained in the manuscript.

## ACKNOWLEDGEMENTS

We thank Dr. Yan Jia for providing assistance with the creation of the sepsis model. This research was partly supported by the National Institute for Health Research (NIHR) Yorkshire and Humber Patient Safety Translational Research Centre (NIHR Yorkshire and Humber PSTRC). The views expressed in this article are those of the authors and not necessarily those of the NHS, the NIHR, or the Department of Health and Social Care.

## CONTRIBUTORSHIP STATEMENT

IH and TL conceived the study. SC, IH, CMC, OJ and MdK planned the study. TL, AK and LF provided clinical expertise for the study. SC led the analysis with assistance from all co-authors. SC drafted the manuscript. All co-authors contributed to the revision of the manuscript and approved the final version.

